# The rich-to-poor vaccine donation game: When will self-interested countries donate their surplus vaccines during pandemics?

**DOI:** 10.1101/2021.12.30.21268537

**Authors:** Adam Lampert, Raanan Sulitzeanu-Kenan, Pieter Vanhuysse, Markus Tepe

## Abstract

When will self-interested vaccine-rich countries voluntarily donate their surplus vaccines to vaccine-poor countries during a pandemic? We develop a game-theoretic approach to address this question. We identify vaccine-rich countries’ optimal surplus donation strategies, and then examine whether these strategies are stable (Nash equilibrium or self-enforcing international agreement). We identify parameter ranges in which full or partial surplus stock donations are optimal for the donor countries. Within a more restrictive parameter region, these optimal strategies are also stable. This implies that, under certain conditions (notably a total amount of surplus vaccines that is sufficiently large), simple coordination can lead to significant donations by strictly self-interested vaccine-rich countries. On the other hand, if the total amount that the countries can donate is small, we expect no contribution from self-interested countries. The results of this analysis provide guidance to policy makers in identifying the circumstances in which coordination efforts are likely to be effective.

## Introduction

The Covid-19 pandemic has shone a harsh light on the urgent need to better understand the drivers of policy solutions to global infectious disease crises. By November 2021, the Covid-19 pandemic had claimed the lives of more than 5.23 million people worldwide ([Covid-19 Dashboard, JHU](https://www.arcgis.com/apps/dashboards/bda7594740fd40299423467b48e9ecf6)), and had led to severe economic losses. During most of 2020 and before effective vaccines and treatments were available, containing the spread of the virus compelled countries to adopt lockdowns and social distancing policies. In a remarkable scientific achievement, safe and effective vaccines were developed in record time (Ghebreyesus 2021; Wouters et al. 2021). The availability of COVID-19 vaccines offers countries and the international community important new means to combat the pandemic. At this particular stage in pandemic cycles, key questions are how best to utilize the available vaccines on a global scale, and what are the strategies that are likely to receive the required international cooperation for their implementation.

Scarcity in supply, coupled with unequal allocation of the available vaccines across countries, present the most formidable challenges for maximizing the potential benefits of these vaccines (Tregoning et al. 2021). Allocating doses internationally in proportion to countries’ population sizes is estimated to be a close-to-optimal strategy in terms of averting deaths worldwide (Hogan et al. 2020) and benefiting the economy, both globally and in donor countries (Çakmaklı et al. 2021; Hafner et al. 2021). Nevertheless, extremely unequal distribution has typified the availability of vaccines across countries (Ghebreyesus 2021, Padma 2021), as depicted in Fig. 1. This situation means that most people in the world’s poorest countries might not have access to COVID-19 vaccines until at least mid-2023 (Padma 2021). Since about 85% of the global population resides in low- and middle-income countries, most of humanity remains exposed to continued outbreaks (Padma 2021). This situation increases the risk that further virus variants will emerge, possibly undermining the efficacy of existing vaccines (Ghebreyesus 2021, Wouters 2021).

**Fig. 1:**
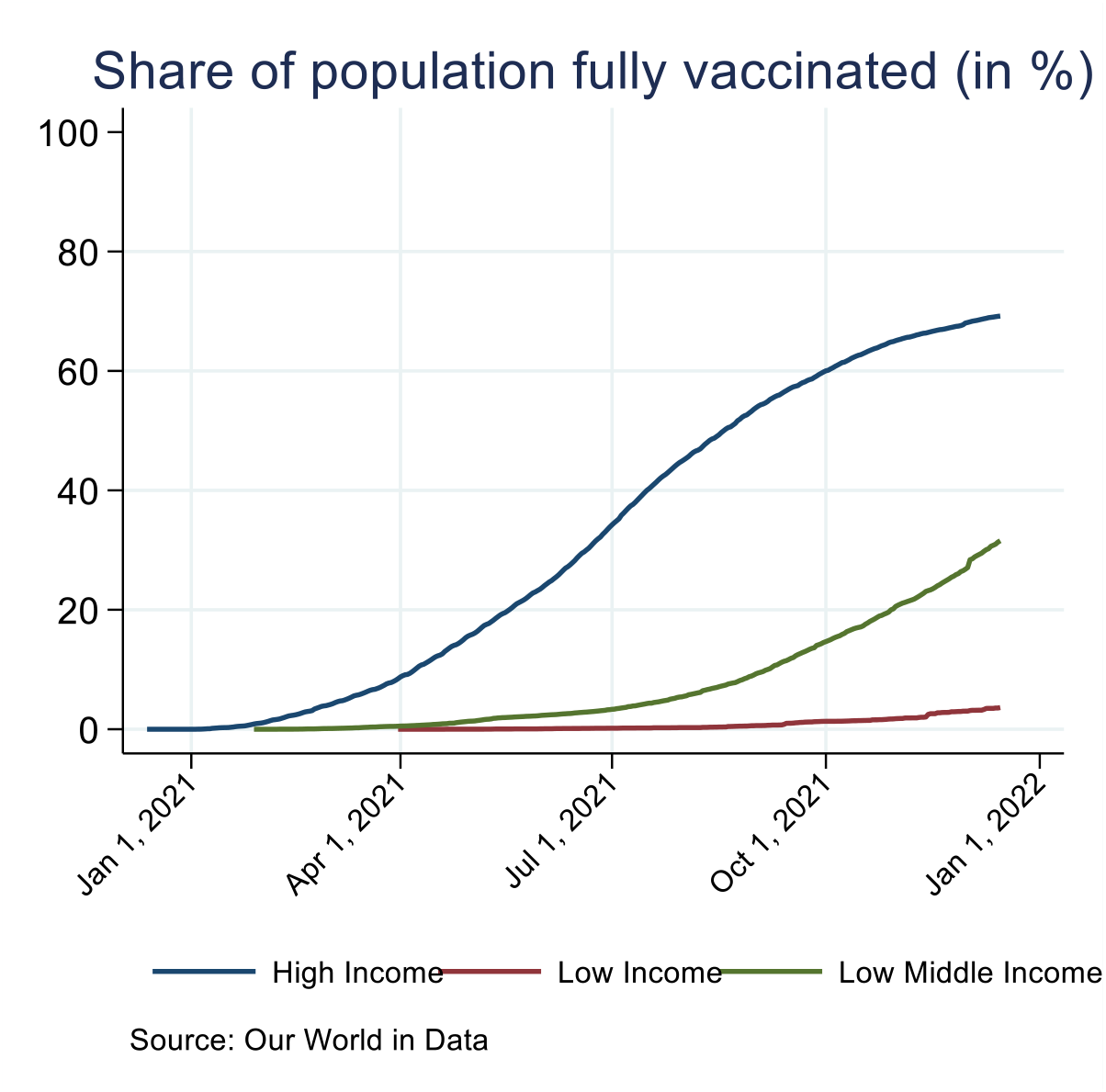
Share of the population fully vaccinated against Covid-19, across high-, lower middle-, and low-income countries.

A notable attempt to distribute vaccines from richer to poorer nations is COVAX - the COVID-19 Vaccine Global Access Facility. Led by the World Health Organization and coordinated jointly with the Coalition for Epidemic Preparedness Innovations and the Global Alliance for Vaccines and Immunization, COVAX is a pooled procurement initiative that aims to provide all countries with COVID-19 vaccines by supplying vaccines at differential prices (Ghebreyesus 2021, Reuters June 13, 2021, Wouters et al. 2021). The guiding idea of COVAX is to prioritize vaccination globally by sub-populations: from older adults, healthcare workers, and other high-risk individuals to the wider sections of the population. It stipulates that no country should vaccinate more than 20% of its population until all countries have vaccinated 20% of their populations (Wouters et al. 2021).

However, the COVAX initiative has so far failed to come close to its objectives^4^, due to the aggregate consequences of vaccine nationalism (Vanhuysse, Jankowski & Tepe 2021). Many of the world’s wealthiest countries have adopted procurement strategies that prioritize widespread inoculation of national populations ahead of the vaccination of health care workers and high-risk populations in poorer countries (Ghebreyesus 2021; Wouters et al. 2021).

The challenges facing globally effective supply of vaccines through supranational initiatives such as COVAX, have led to initiatives that focus on country-level and ad hoc intergovernmental approaches, such as donations by high-income countries of some of their pre-purchased vaccines to medium and low-income countries (Clarke et al. 2021). The G7 summit of June 2021 jointly pledged to provide 1 billion COVID-19 vaccine doses until June 2022 (Reuters June 13, 2021). Such initiatives focus on ways to redistribute the excess stocks of doses accumulated by vaccine-rich countries *after* they have vaccinated large shares of their own populations. They echo similar attempts in previous global health crises, such as smallpox in the 1970s (Barrett 2007), HIV in the 1980s (Ghebreyesus 2021), and the 2009 H1N1 (Wouters et al. 2021).

This study adopts a game-theoretic approach to identify the conditions under which self-interested vaccine-rich countries would donate their surplus vaccine doses (beyond those needed for the initial vaccination of their own population) to vaccine-poor nations, rather than stocking these surplus doses domestically for future own-nation use.^5^ Vaccine-rich countries have at least two self-interested reasons for sharing their vaccine surpluses in this way. First, they have large open economies that are dependent on international trade and thus require significant levels of international travel and open borders. Second, their efforts to stop the pandemic within national borders could be undermined by the emergence of new variants of concern (VOCs). Such a VOC (e.g., the Delta and Omicron variants) may require increasing the level of immunization in the population by administering a booster (Rosenberg et al. 2021). In such circumstances, having a stock can significantly shorten time-to-delivery, thereby containing the outbreak and saving domestic lives. The probability of a VOC is a crucial parameter to understand vaccine-rich governments’ willingness to re-allocate vaccines to vaccine-poor countries. The fear of VOC can either increase short-term national self-interest and a tendency to reserve health resources for domestic purposes or create awareness that long-term pandemic control can only succeed by effective global vaccination. Another complicating element is that donation by one country benefits all others by reducing the probability of future variants of the virus and their consequent costs; however, such a donation incurs a cost for the donating country by leaving it without an available stock in the case of a VOC outbreak.

This study models (1) what is the vaccine donation strategy that is optimal for all vaccine-rich countries combined (a social planner’s perspective); and (2) whether the optimal solutions could be adopted by the relevant countries, assuming strictly self-interested motivation on their part. In particular, we examine how the answers to these questions depend on key pandemic parameters: (1) the fraction of the global unvaccinated population potentially covered if all vaccine-rich countries fully donate their surplus (*v*_max_); (2) the baseline expected annual rate of VOCs (λ); and (3) the fraction of the total cost of a new VOC outbreak that is unavoidable despite having surplus doses (*α*). The game-theoretic model we develop is general in the sense that it identifies the conditions under which a minority of vaccine-rich countries are likely to donate a costly remedy in the context of a pandemic. By considering the realistic ranges of the model parameters in the case of Covid-19, we can cautiously infer more specific implications for the potential of international cooperation in coping with this particular pandemic.

## Model

### The rich-to-poor vaccine donation game

We consider the strategies of *N* vaccine-rich countries (hereafter: ‘the countries’). Beyond the vaccine doses needed to vaccinate their own entire population, each vaccine-rich country is assumed to have purchased an additional stock sufficient to vaccinate its population two *additional* times. The benefit of having this surplus full-population stock is that it could be administered in case a VOC emerges.^6^ Each country can decide how much out of the two extra doses per capita it donates to vaccine-poor countries: *s*_*i*_ = 0,1, or 2, where *i* = 1,2,3, … *N* denotes the country. Each country can donate zero extra doses and thus keep two extra doses per capita for itself; donate one dose and keep 1 for itself; or donate both extra doses and keep none (1 dose may protect against 1 variant; 2 doses may protect against 2 variants.)

We denote *v* as the fraction of the global unvaccinated population that could be vaccinated due to the vaccine-rich countries’ donations.^7^ It could vary from *v* = 0 if no country donates (*s*_*i*_ = 0 for all *i*) to *v*_max_ if all the countries donate both doses (*s*_*i*_ = 2 for all *i*). Specifically, if *v*_max_ ≥ 1, the countries could vaccinate the entire unvaccinated world population and be fully protected against any variants. However, if *v*_max_ < 1, there is still a chance that variants will emerge even if all the countries contribute all their extra doses.

In turn, we denote *λ* as the expected number of VOCs that emerge in unvaccinated populations within one year, if no doses are donated. We assume that variants can emerge independently with some small probability in each unvaccinated person (Fontanet et al. 2021). Therefore, if vaccines are donated, the expected annual rate of VOCs is given by *λ*(1 − *v*) if *v* ≤ 1 and 0 if *v* > 1. Specifically, if *v* < 1, the actual number of VOCs that emerge is given by a Poisson distribution with a mean *λ*(1 − *v*). It follows that the probability for the occurrence of exactly *k* variants, *P*_*k*_, is given by the Poisson coefficient

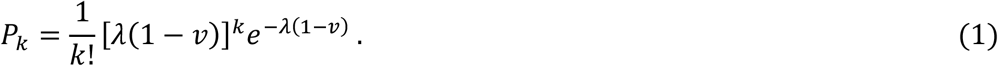

Donation by country *i* benefits all countries because it reduces the probability of future virus variants and their consequent costs. However, a donation also comes with a cost to the donor country because it might be left without an effective vaccine if variants do occur. Specifically, if a country stocked enough doses to cover the occurrence of a variant, it will have to bear only a fraction *α* (0 < *α* ≤ 1) of the total cost of the corresponding outbreak. Note that *α* is expected to be greater than zero – i.e., a certain cost of a future outbreak is practically unavoidable – since some time is required for administering surplus vaccines to the population.^8^ During this interim period, the country is expected to bear the cost of the variant outbreak. Therefore, the expected cost of future outbreaks to country *i* is given by

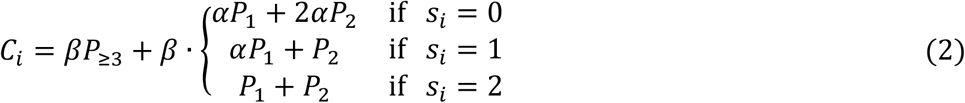

where *β* is the cost due to an outbreak if the country does not have extra vaccine doses, and *P*_≥3_ is the probability that three or more variants emerge. Note that *P*_1_, *P*_2_ and *P*_≥3_ depend on *v* (Eq. 1), therefore they depend on *s*_*i*_ and on the strategies of all other countries.

### Analysis of the model: two solution concepts

Using this model, we address the following two tasks. First, we identify the optimal solutions from the point of view of a ‘rich-world’ social planner who aims to maximize the social welfare of all vaccine-rich countries combined. Specifically, this optimal solution is given by the choice of *s*_*i*_ for each *i* that minimizes the total expected cost of outbreaks to all the vaccine-rich countries:

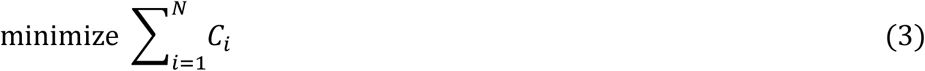

After identifying the optimal solution, we ask whether this solution is stable. Assuming that each rich country chooses the strategy *s*_*i*_ that minimizes its own expected cost, will the optimal solution be adopted? To answer this question, we consider two solution concepts. The first one is the Nash equilibrium. Assume, for example, that *s*_*i*_ = 2 for each country is the optimal solution. This is also a Nash equilibrium if and only if it does not benefit any of the countries to unilaterally change its strategy to *s*_*i*_ = 1 or *s*_*i*_ = 0. However, the Nash equilibrium solution concept is somewhat restrictive, because it does not take into account the response of the other countries to the deviation of a given country.

Another widely used solution concept in the context of international cooperation is a self-enforcing international agreement (henceforth SEA) (Barrett 1994). The assumption is that each country chooses whether to be a signatory or not. The non-signatories do not contribute (or contribute less), while the signatories adopt the strategy that maximizes their welfare (minimizes their expected cost) as a whole. An agreement is self-enforcing if no signatory has an incentive to opt-out and become a non-signatory, and no non-signatory has an incentive to opt-in and become a signatory (Barrett 1994). Note that if the optimal solution is a Nash equilibrium, it is also a SEA. However, there could be cases in which the optimal solution is a SEA but not a Nash equilibrium. For example, in the context of our game, consider the case in which *s*_*i*_ = 2 by all countries is not a Nash equilibrium: if all countries adopt *s*_*i*_ = 2, it may benefit country *j* to deviate to, say, *s*_*j*_ = 1. However, following this opting out by country *j*, the best strategy of the remaining countries (the signatories) may become *s*_*i*_ = 1 instead of *s*_*i*_ = 2. If country *j* anticipates that the remaining countries will reduce their contribution to *s*_*i*_ = 1 in response to its own deviation, it might be beneficial for country *j* to persist with *s*_*i*_ = 2.

## Results

### Optimal solution

We first examine what the optimal solution is from the point of view of a social planner who aims to maximize the social welfare of all vaccine-rich countries combined. Specifically, we find the contribution that maximizes the social welfare (minimizes the expected cost) of the *N* vaccine-rich countries as a whole across the various parameter ranges. Due to the substantial uncertainty regarding some parameter values, such as the expected rate of VOC (*λ*), the analysis considers the following parameter ranges: 2 ≤ *N* ≤ 10, 0 ≤ *α* ≤ 0.5, 0 ≤ *v*_max_ ≤ 1, and 0 < *λ* ≤ 2.^9^ The top panel in Fig. 2 maps the optimal solutions and their stability to parameters *v*_max_ (Y-axis) and *λ* (X-axis), holding the number of countries and effectiveness of stocking constant (*N*=6 and *α*=0.4, respectively). In turn, each panel in Figs. 3 and 4 also shows the optimal solutions and their stability as a function of *α* and *N*.

**Fig. 2:**
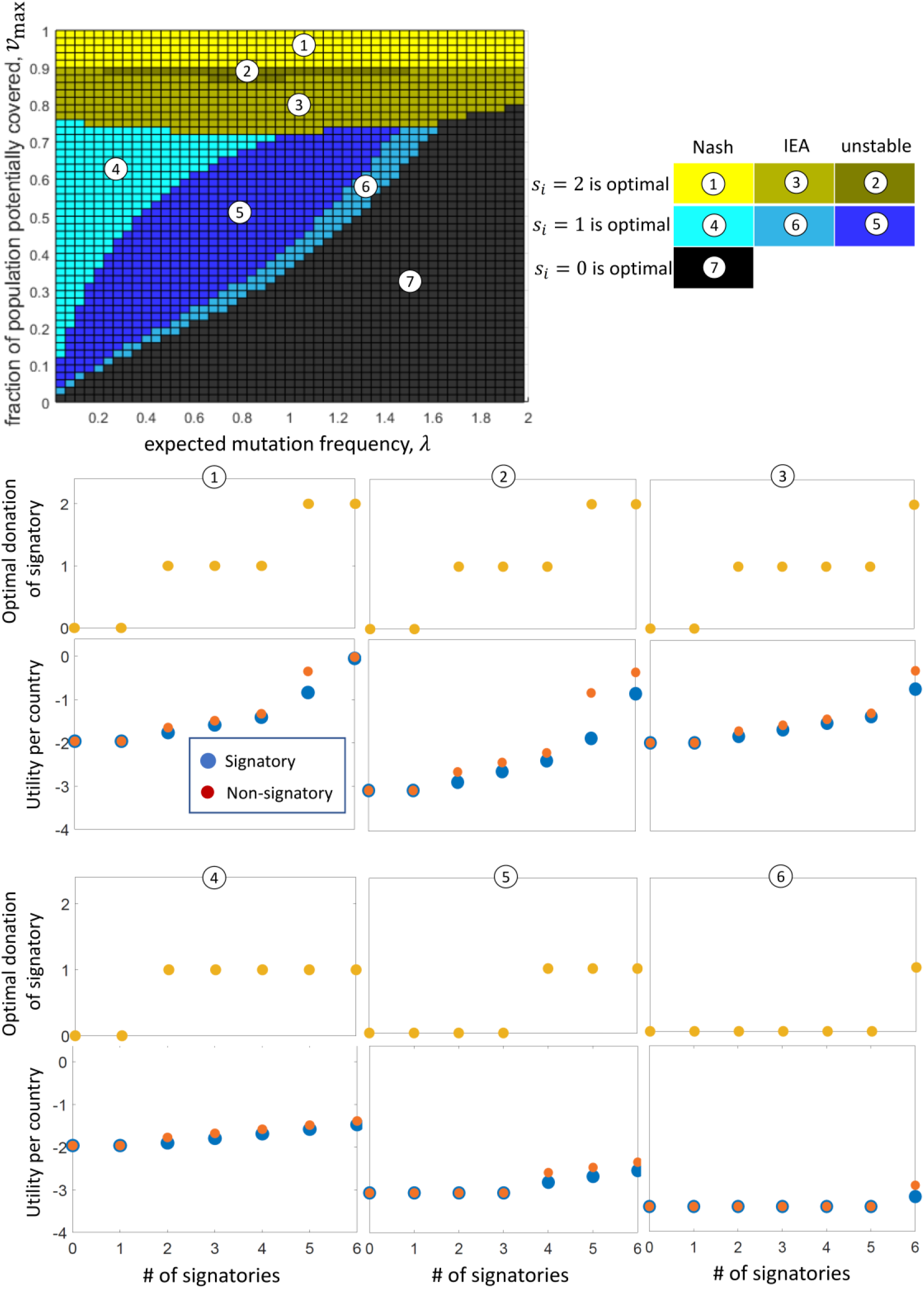
The optimal solution and its stability depend on the values of the parameters *v*_*max*_ and *λ*. The optimal solution is given by the strategies *s*_*i*_ that minimize the total expected cost to all vaccine-rich countries as a whole (Eq. (3)). Each colored rectangle on the top panel represents the solution for the corresponding values of *λ* and *v*_max_, where the other parameters are *α* = 0.4 and *N* = 6. For some parameter regions, denoted as regions 1, 2, and 3 (yellow shades), *s*_*i*_ = 2 by all rich countries is optimal. For some other parameter regions, denoted as regions 4, 5, and 6 (blue shades), *s*_*i*_ = 1 by all countries is optimal. Otherwise (region 7, black), *s*_*i*_ = 0 by all countries is optimal. The parameters also determine whether the optimal solution is stable and can be adopted by rational agents, where each country acts self-interestedly. In regions 1 and 4, the optimal solution is also a Nash equilibrium. The panels corresponding to regions 1 and 4 demonstrate that the welfare (−*C*_*i*_) for a country that adopts the optimal solution is larger than that adopting *s*_*i*_ = 0 (blue dot in the case of 6 signatories is higher than the red dot for 5 signatories). In turn, in regions 3 and 6, the solution is not a Nash equilibrium but is a self-enforcing international agreement (SEA). (Note that if the optimal solution is a Nash equilibrium, it is also a SEA.)

**Fig. 3:**
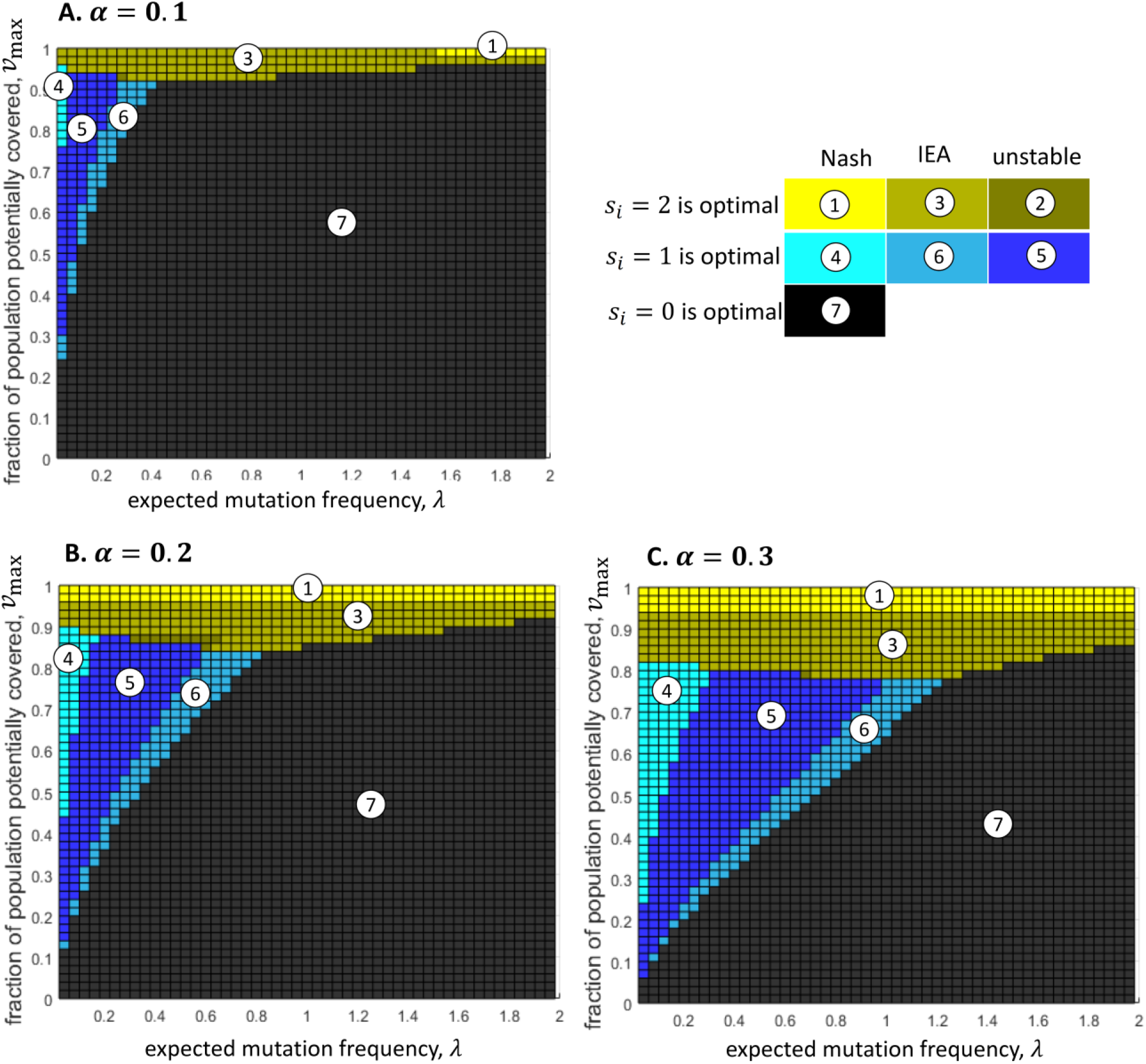
The optimal solution and its stability depend on the values of the parameters *v*_*max*_, *λ*, and *α*. Each panel characterizes a different value of *α*, where *α* = 0.1 in panel A, *α* = 0.2 in panel B, and *α* = 0.3 in panel C (*N* = 6 in all panels). One notable observation is that if *α* (the unavoidable fraction of the cost of an outbreak, given surplus doses) is smaller, then it is less worthwhile to donate, and the range of parameters that renders donations advantageous is narrower.

**Fig. 4:**
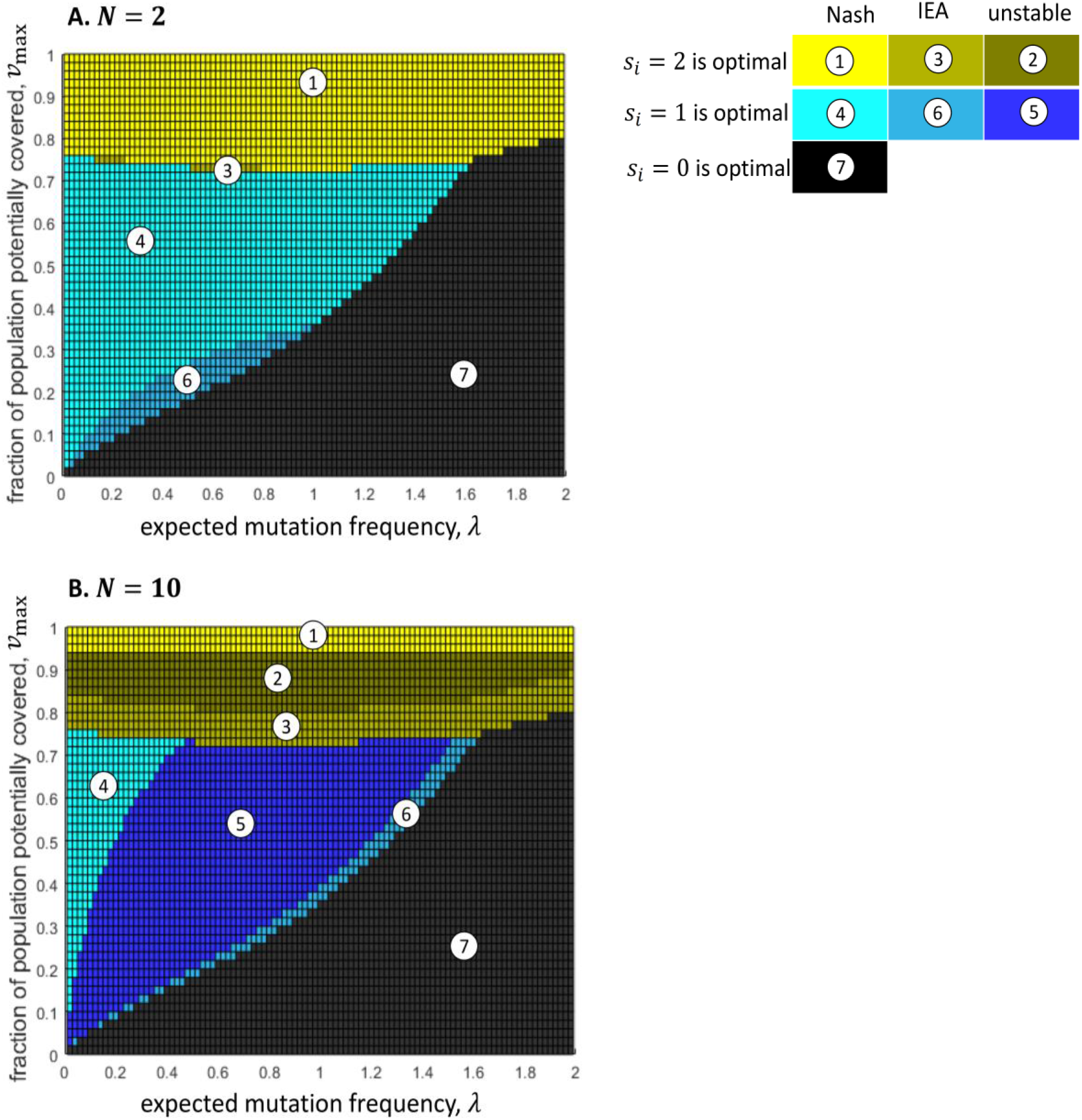
The optimal solution does not depend on the number of rich countries, *N*, as long as *v*_max_ is held constant; however, the optimal solution is stable for a wider range of parameters if *N* is smaller. The only difference in parameters between the panels is that *N* = 2 in panel A and *N* = 10 in panel B (*α* = 0.4 in both panels). The case of *N* = 6 is demonstrated in Fig. 2.

Our results show that the optimal solution is always an equal donation by all countries, of either 0, 1, or 2 full-population rounds of vaccinations, depending on the values of the parameters *λ, α, N*, and *v*_max_ (Figs. 2-4).^10^ Specifically, zero donation of surplus vaccines by all countries (*s*_*i*_ = 0 for all *i*) is optimal in parameter regions where *λ* is sufficiently large and *v*_max_ and *α* are sufficiently small (black region 7 in Figs. 2-4). Full donation (*s*_*i*_ = 2 for all *i*) is optimal if *v*_max_ is sufficiently close to 1 and/or *α* is sufficiently large (yellow and yellow-grey regions 1, 2, and 3 in Figs. 2-4). In turn, donation of half of the surplus vaccines (*s*_*i*_ = 1 for all *i*) is optimal if *λ* is sufficiently small, and *v*_max_ and *α* have some intermediate values (blue and blue-grey region 4, 5, and 6 in Figs. 2-4).

Unsurprisingly, vaccine-rich countries benefit more from donating, the larger *v*_max_ is. If *v*_max_ is much below 1, then even if all the extra vaccine doses are donated, only a small portion of the world’s unvaccinated population can be covered, and the risk of future variants cannot be significantly reduced. On the other hand, if *v*_max_ is close to 1, nearly the entire unvaccinated world population can get vaccinated, thus donating countries can expect to reduce the risk of variants considerably. Interestingly, however, our results indicate that the threshold of *v*_max_ where the optimal strategy becomes *s*_*i*_ = 2, is not very sensitive to *λ* – as shown, for example, in Fig. 2, where *v*_max_ > .75.

Below this threshold of *v*_max_, *λ* acquires a substantial effect on whether *s*_*i*_ = 1 or *s*_*i*_ = 0 is optimal. This is relevant from a policy perspective as it means that in quite realistic ranges of *v*_max_, whether it is beneficial for the vaccine-rich countries to donate an extra dose per-capita or not donate at all depends to a large extent on the expected frequency of VOCs. Specifically, if *λ* is small, it may be beneficial for vaccine-rich countries to contribute one dose per capita each, even under relatively small values of α and *v*_max_ – as depicted by the blue regions (4, 5 & 6) in Fig. 2. By contrast, if *λ* is large, donation is beneficial only if the value of *v*_max_ is larger.

These patterns are modified, yet retain their basic structure, when the unavoidable fraction of the cost of a future outbreak (*α*) varies. Vaccine-rich countries are clearly better off internationally donating more surplus vaccines when *α* rises. In the extreme case, if the time it takes to administer pre-stocked extra doses is long enough to incur the total cost of a new outbreak (*α* = 1), then stocking extra doses domestically is practically worthless, and a better strategy is to donate all surplus doses to the vaccine-poor world to reduce the risk of variants. However, as *α* decreases, it becomes more worthwhile to hold enough doses for an extra vaccination round domestically. Fig. 3 illustrates this with *α* values of respectively 0.1, 0.2, and 0.3. Not surprisingly, the surfaces of the two-dose-optimal regions (1-3) and the one-dose-optimal regions (4-6) are smaller for lower *α* values. Note, lastly, that it follows directly from the model that the optimal solution does not depend on the number of rich countries, *N*, as long as *v*_max_ is held constant, as shown in Fig. 4.

### Stability of the optimal solution: Nash equilibrium and self-enforcing international agreement

Having identified optimal strategies, we next examine whether one could expect the *N* vaccine-rich countries to actually adopt these optimal strategies, acting strictly self-interestedly and voluntarily. Again, the answer depends on the parameter values for *λ, α, v*_max_, and *N* (Figs. 2-4).

*Full donation:* In some parameter regions (denoted as region 1 in Figs. 2-4), donating both surplus vaccine doses is not only optimal but is also a Nash equilibrium: if *N* − 1 countries adopt *s*_*i*_ = 2, then the best response of the last country is also *s*_*i*_ = 2. In that case, we expect the *N* countries to willingly contribute both their surplus full-population stocks to vaccine-poor countries. In other words, under configurations of high values of *v*_max_ and realistic values of *α* (see supplementary information), double-dose donation by vaccine-rich countries is a Nash equilibrium.

Furthermore, in certain parameter regions in which donating *s*_*i*_ = 2 doses is optimal but not a Nash equilibrium, there still remain realistic prospects for stable donation strategies by vaccine-rich countries, as the optimal solution may be a self-enforcing international agreement (SEA). The *N*^*th*^ country might not be willing to deviate from an agreement to donate *s*_*i*_ = 2 if it foresees that the optimal contribution by the remaining *N* − 1 countries will become *s*_*i*_ = 1 (region 3 in Fig. 2). This idea is demonstrated in the bottom panel of Fig. 2. If the 6th country opts out, the other 5 countries will now be better off adopting a strategy in which *s*_*i*_ equals 1 rather than 2 (yellow dot for 5 signatories is lower than for 6 signatories), rendering the utility for the putative non-signatory (6^th^ country) lower than its utility had it not opted-out of the agreement (the red dot with 5 signatories is lower than the blue dot with 6 signatories). In other words, in addition to being a Nash equilibrium, under a still larger set of configurations of realistic values of both *α* and *v*_*max*_, a double-dose donation can also be a SEA.

*Partial donation:* Similarly, for some parameter values (the light blue regions 4 and 6 in Figs. 2-4), donating one dose is both optimal and stable. Specifically, *s*_*i*_ = 1 by all the countries is a Nash equilibrium in region 4. In addition, if *s*_*i*_ = 1 is optimal but not a Nash equilibrium, *s*_*i*_ = 1 by all countries is self-enforcing in parameter region 6.^11^

As Fig. 4 shows, the optimal solution is stable for a wider range of parameters if the number of vaccine-rich countries, *N*, is smaller. Increasing *N* from 2 to 10, while holding the other parameter values constant, diminishes the set of parameter configurations in which both *s*_*i*_ = 2 and *s*_*i*_ = 1 are a Nash equilibrium (regions 1 and 4, respectively), as depicted in Fig. 4. The reduction in the set configurations in which donations are a stable strategy as *N* increases is because the benefit to a given country due to its own contribution becomes smaller, while the cost per capita to the donating country remains the same.

Finally, note that in other regions, donation of two doses (region 2) or one dose (region 5) is optimal (maximizes the social welfare of the vaccine-rich countries as a whole), but the optimal solution is unstable. Instead, the stable solutions in these parameter regions, both Nash equilibria and SEAs, are sub-optimal and dictate either no donation, donation by only some countries, or donation of only one dose when the optimum is two doses. The exact solution depends on the parameter values (within regions 2 and 5). For simplicity, Figs. 2-4 only indicate the parameter ranges in which the optimal solution is stable (regions 1, 3, 4, and 6) or unstable (regions 2 and 5), without indicating the stable solutions for every parameter value within the unstable regions, and without indicating other possible Nash equilibria in regions where the optimal solution is stable.

## Discussion, limitations and implications

This study adopts a game-theoretic approach to explore the conditions under which strictly self-interested vaccine-rich countries would voluntarily donate their surplus vaccine doses to vaccine-poor countries in the context of a pandemic, without recourse to any other motivations such as international solidarity. We addressed this question by first identifying the optimal vaccine donation strategies for vaccine-rich countries and then examining whether these optimal strategies could be adopted by these vaccine-rich countries, assuming that they act as strictly self-interested, rational agents.

We show that full donations are optimal when their potential impact is high (*v*_max_), stocking surplus doses is less effective in averting the cost of a future outbreak (*α*). Specifically, if the fraction of the unvaccinated world population that can be vaccinated if all vaccine-rich countries fully donate (*v*_max_) is above a certain threshold, full donation is optimal. The value of this threshold of *v*_max_ is determined by *α*. Interestingly, above this threshold the optimal solution of full donations is not sensitive to the expected annual rate of VOCs (*λ*). Below this threshold, partial donations or no donations are optimal, depending jointly on the rate of VOCs (*λ*), *v*_*max*_, and the unavoidable fraction (*α*) of the cost of future outbreaks (despite stocking surplus doses). These results point to the importance of *v*_max_. Although the expected number of variants ex-ante affects the utility of donations, it is also ex-post influenced by donations, depending on their potential impact on the unvaccinated world population. If *v*_max_ is sufficiently large, the pre-donation rate of expected variants becomes inconsequential for the utility of donations, as reflected by the results. However, if *v*_max_ is relatively low, pre-donation rate of expected variants is an important determinant of the utility of vaccine donation.

This study’s second and crucial finding is that in certain parameter regions, self-interested vaccine-rich countries can be expected to willingly donate the socially optimal surplus vaccine doses. That is, we show that these optimal surplus donation strategies are also stable. Specifically, when *full* (double-dose) donation is optimal, it is also a Nash equilibrium if *v*_max_ is sufficiently large (region 1 in Figs. 2-4), and it is a self-enforcing agreement in a somewhat lower range of *v*_max_ (region 3). When *partial* (single-dose) donation is optimal, it is also a Nash equilibrium if the risk of future variants of concern (*λ*) is relatively low and *v*_max_ is at medium levels (region 4), and it is a self-enforcing agreement in a narrower set of parameters regions (region 6). Lastly, while the number of potential donor countries (*N*) does not affect the optimal solution, a larger number of countries does imply a narrower range of parameter values in which the optimal solution is stable (Fig. 4). Thus, if the aggregate stock of surplus doses of vaccine-rich countries is distributed over a large number of such countries, donations are less likely to be a stable strategy even when they are optimal.

Tentative policy implications can be inferred from these findings. Our results offer hope for global effective pandemic solutions as they identify significant windows for self-interested rich-to-poor world vaccine donation. That said, we also see large parameter ranges where the prediction is zero donation. Given our assumption that donations are feasible only when they are the optimal solution, within the parameter ranges where this is the case, coordination efforts are likely to be fruitful. The results of this study may contribute to the efforts of organizations such as the World Health Organization in promoting such endeavors. In the supplementary information, we provide estimates for our model parameters in the context of the current Covid-19 pandemic.

The vaccine donation game presented here offers greater potential for international cooperation, compared, for example to climate agreements, due to the positive feedback between the marginal benefit from donation and the amount donated by other countries. Specifically, if a given country donates, it decreases the number of unvaccinated people worldwide, which makes the contribution of any other country more valuable because its donation now covers a greater portion of the unvaccinated world’s population. Therefore, the contribution of a given country increases the incentive of the other countries to contribute. This is fundamentally different from the case of international climate agreements (Barrett 1994; Finus 2008), in which the contribution of some countries makes it less beneficial for more countries to join a treaty, because climate cost is assumed to increase super-linearly with temperature changes.

The literature on vaccine donation games goes back to Barrett (2007), who examined the international subsidy of Smallpox vaccine in poor countries. That problem was, however, fundamentally different from ours and characterized a much later stage of the pandemic. There was no question about the optimality of the subsidy and there was no shortage of vaccine dosage; the question was which country would be the one that invests the money (Barrett 2007). In another noteworthy study, Lampert (2021) studies the international subsidy for treatment of harmful invasive species in poor countries to prevent their spread into rich countries. He showed that, in Nash equilibrium, fewer countries might do a better job than multiple countries. In that study, however, a single or a few rich countries had the capacity of eliminating the species even when working alone. Finally, in the context of the Covid-19 pandemic, Caparrós and Finus (2020) studied the influence of countries’ policies on each other, focusing on pre-vaccine Covid-19 preventative measures. They concluded that the game is analogous to a weakest-link game: a few countries that do not take sufficient preventative measures are sufficient for spreading the pandemic internationally. Note that our vaccine donation game is fundamentally different and does not necessitate the contribution of all countries.

Our analysis is intended to offer a general game-theoretic approach to solving the problem of donating surplus vaccines during a pandemic. It unavoidably incorporates several simplifying assumptions. First, we assume that countries stock excess vaccine doses for the purpose of responding quickly to an outbreak that is due to a VOC. However, countries also stock vaccines due to uncertainty regarding the duration of vaccine efficacy, or in order to diversify the stock of vaccines (by producers and technologies) due to uncertainty regarding their consequent approval and efficacy. These additional reasons for stocking surplus vaccine doses are not related to the risk of VOC, thus to the extent that they are dominant, may decrease the likelihood of donations.

Stocking vaccine doses in practice may be implemented in several ways. It can be done through the actual stocking of vaccine vials within the country’s territory, but this method is often limited due to vaccines’ storage periods. A second method is to obtain purchasing contracts that prioritize the supply of additional vaccines, if required. Donations are possible in all three methods. However, the different methods entail varying response times, in the event of an outbreak, with the potentially quickest response in the case of physical stocks, and longest in the case of a dedicated budget. These different response durations are captured in our model by *α*, thus specific information about the stocking methods of each vaccine-rich countries can be used to enhance the estimates in concrete circumstances.

Lastly, vaccine-rich countries in our model are assumed to be equal in size. Relaxing this assumption may change some of the results. Notably, it is likely that the optimal strategy will not be the same for the entire set of rich countries. Moreover, variation in vaccine-rich country sizes may alter the results regarding solution stability. Further research is required in order to apply the model to real-life differences between vaccine-rich countries.

## Data Availability

All data produced in the present work are contained in the manuscript

## Methods

### Calculating the optimal solution

To calculate the expected cost to a given country *i, C*_*i*_, given the set of strategies that all the countries adopt, we first calculate *P*_1_ and *P*_2_ (Eq. 1), and then we set *P*_≥3_ = 1 − *P*_1_ − *P*_2_ and substitute the results in Eq. (2). In turn, we examine all possible sets of strategies, where in each set, each country can contribute a different amount (*s*_*i*_ equals 0, 1, or 2 for each *i*). For each set, we calculate the total cost (Eq. (3)), and we find the set for which the total cost is minimized.

### Checking whether the optimal solution is a Nash equilibrium

Without loss of generality, we examine whether country 1 can benefit from deviating if all the countries adopt the optimal solution, *s*_*i*_ = *s*^opt^. Specifically, country 1 has 2 possible deviations: to *s*_1_ = 0 or *s*_1_ = 1 if *s*^opt^ = 2, and to *s*_1_ = 0 or *s*_1_ = 2 if *s*^opt^ = 1. We first calculate 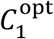, the expected cost to country 1 if it does not deviate (and the other countries adopt *s*_*i*_ = *s*^opt^). Then, we calculate 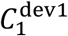 and 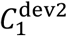, the expected cost to country 1 following each of its possible deviations. We conclude that the opt solution is a Nash equilibrium if and only if 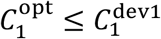 and 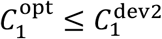.

### Checking whether the optimal solution is a self-enforcing international agreement

For every 1 ≤ *n* ≤ *N*, we consider *n* signatories and *N* − *n* non-signatories, and we calculate the utility (minus the cost – Eq. (2)) of a signatory, *u*_*s*_(*n*), and the utility of a non-signatory, *u*_*o*_(*n*). (Note that, for each *n, u*_*o*_(*n*) ≥ *u*_*s*_(*n*) (Barrett 1994)). In turn, an agreement with *n* = *N* signatories is, by definition, the optimal solution. This solution is a self-enforcing international agreement if a country cannot benefit from opting-out when all *N* countries are signatories. Specifically, if a country remains a signatory, its utility is *u*_*s*_(*N*), while if it opts-out, its utility becomes *u*_*o*_(*N* − 1). Therefore, the optimal solution is self-enforcing if and only if *u*_*s*_(*N*) ≥ *u*_*o*_(*N* − 1).

## Supplementary information

### Estimating the parameter values in the context of the Covid-19 pandemic

#### Estimating *v*_max_ in the context of the Covid-19 pandemic

Given a world population of 7.9 billion (as of Oct. 2021); the share of vaccinated globally is .4216 (fully vaccinated) and .1139 (partially vaccinated), we estimate the global proportion of vaccinated population at .4786 (.4216+.1139/2).^12^ Thus the number of unvaccinated is estimated at 4.119 billion (.52145×7.9).

Based on this estimate and the on the number of ordered vaccine doses around the world (missing data for Russia and Turkmenistan)^13^, we can provide some estimates for *v*_max_. If we order countries by the size of their surplus doses stock, the first two political entities – the European Union and the United States) – offer *v*_max_ = .53; and the first ten provide *v*_max_ = .85. It should be noted that the size of surplus doses vary between countries, while our theoretical model assumes that vaccine-rich countries are equal.

#### Estimating *α*

We assume that a country is only required to re-vaccinate its population in the advent of a new outbreak due to a VOC. One way to approximate *α* is to estimate the time it takes to administer the vaccine to a sizable proportion of the population (50%), vis-à-vis the length of an outbreak wave (absent a vaccine). Based on the duration of the second outbreaks of Covid-19 in 12 countries (Covid-19 Data Explorer), a typical outbreak lasts 5.9 months (SD=1.83), but we assume its costs (health implications and economic disruption) linger further up to 10 months. If a country has vaccines available to provide a booster shot, it will incur the cost of the outbreaks until it manages to re-vaccinate a sizable proportion of its population. The average time it took the five vaccine-rich countries (table below) to reach this level of vaccination during 2021 is 6.19 months. However, in some of these countries delays in vaccinations were due to vaccine shortages (see: New-York Times Jan. 27 2021). Thus, assuming no shortages and given that a booster shot requires just one dose, this duration may be only 3-4 months, yielding a plausible range of 0.3 < *α* < 0.6.

**Table A1:** Durations of public vaccination^14^

|  | Time to 50% fully vaccinate<br>(in months) |
| --- | --- |
| Canada | 6.72<br>(12.12.2020-7.19.2021) |
| France | 6.69<br>(1.16.2021-8.7.2021) |
| Germany | 7.00<br>(12.27.2020-7.27.2021) |
| Israel | 3.10<br>[12.19.2020-3.24.2021] |
| United Kingdom | 5.90<br>(1.10.2021-7.7.2021) |
| United States | 7.77<br>(12.13.2020-8.6.2021) |
| Average | 6.20 [95% CI: 4.48-7.9] |

#### Estimating ***λ***

As noted in the main text, the utility of stocking surplus vaccine doses is associated with the likelihood of VOCs. The rate of VOC emergence is, to the best of our knowledge, unknown. A tentative assessment of the two year experience with known Covid-19 VOCs suggests that, while there have been infections with VOCs among vaccinated, vaccine efficacy in reducing the severity of disease was retained for the first four VOCs (Tregoning et al. 2021). However, at present, it is still unknown whether the fifth VOC reported recently by the WHO (B.1.1.529, Omicron) threatens vaccine effectiveness (Callaway 2021). Given this evidence, we cautiously maintain that a plausible range for the rate of VOCs per year is 0.5-1.5. To the extent that more accurate estimates are available, either in the context of the current pandemic or the next, we considered an even wider range of 0 ≤ *λ* ≤ 2.

## Footnotes

4 COVAX will not reach its pledge to deliver two billion doses by the end of 2021, as by mid-October only 288 million doses had been delivered to low and middle income countries (Usher 2021).

5 Note that stocking in practice might be based on pre-purchased doses that can be supplied on a short notice from the manufacturer. These contractual arrangements are required to avoid expiration of stored vaccine vials (Interview with Roee Reicher, Head of health sector, budget department, Israeli Ministry of Finance, Nov. 4, 2021).

6 Another reason to stock extra doses is potential waning of the current vaccine’s protection against the existing strand of the virus. However, in this study we concentrate on the risk of an outbreak due to a new variant of concern (VOC). The two risks (waning protection and emergence of a VOC) are qualitatively interchangeable as both present a risk regarding the future efficacy of the existing vaccine.

7 Given the assumption that countries will donate only after fully vaccinating their domestic population, “global unvaccinated population” refers to the unvaccinated in vaccine-poor countries. In the Discussion section we consider the reality of imperfect immunization in vaccine-rich countries due to vaccine hesitancy (Machingaidze and Wiysonge 2021).

8 For example, if a booster shot is required, the time and direct costs of administering the booster shot remain. Note that α may, more generally, capture the time/cost required to apply any stocked medial resource.

9 Parameter ranges were chosen based on estimates of their plausible values in the context of the Covid-19 pandemic (see: supplementary information). However, the model is not limited to these specific parameter ranges.

10 Note that it is not trivial that the optimum solutions do not include one in which different countries choose different strategies. This result follows as there is a positive feedback between the marginal benefit from donation and the amount donated by other countries; therefore, if it is beneficial for the countries to increase their donation from *s*_*i*_ = 0 to *s*_*i*_ = 1, then it is even more beneficial to increase the donation if some countries already donate.

11 Note that SEAs that are not a Nash equilibrium appear in parameter values that are very close to those where the optimal strategy becomes *s*_*i*_ = 1 instead of *s*_*i*_ = 2 (region 3), or becomes *s*_*i*_ = 0 instead of *s*_*i*_ = 1 (region 6). The reason is that, in those regions, the deviation of one country can change the optimal strategy of the remaining *N* − 1 countries (see lower panels in Fig. 1).

12 Source: https://ourworldindata.org/covid-vaccinations. Data extracted on November 24, 2021.

13 Source: https://www.economist.com/graphic-detail/tracking-coronavirus-across-the-world. Data extracted on October 26, 2021. This measure links our estimates to existing production capacity, rather than merely the willingness of countries to invest in vaccinations. Since these data exclude stocking in the form of a dedicated budgets, our analysis here yields conservative estimate for both v_max_.

14 Source: https://ourworldindata.org/covid-vaccinations. Data extracted on November 24, 2021.

